# Management of Nontuberculous Mycobacterial Pulmonary Disease Refractory to Guideline-Based Therapy: A Systematic Review

**DOI:** 10.1101/2025.06.23.25330100

**Authors:** Diana Moreira-Sousa, Beatriz Martins, Ana Aguiar, Marina Pinheiro, Onno W. Akkerman, Timothy R Aksamit, Stefano Aliberti, Claire Andrejak, Charles L. Daley, Jakko van Ingen, Christoph Lange, Marc Lipman, Michael R Loebinger, Mateja Jankovic Makek, Kozo Morimoto, Rachel Thomson, Dirk Wagner, Kevin L Winthrop, Jae-Joon Yim, Raquel Duarte

**Affiliations:** Pulmonology Department, Local Health Unit Cova da Beira, Covilhã, Portugal; Pulmonology Department, Local Health Unit São João, Porto, Portugal; EPIUnit ITR, Instituto de Saúde Pública da Universidade do Porto, Universidade do Porto, Rua das Taipas, n° 135, 4050-600 Porto, Portugal, Estudo das Populações - Instituto de Ciências Biomédicas Abel Salazar, Universidade do Porto, Porto, Portugal; Public Health Unit, Local Health Unit Barcelos/Esposende, Barcelos, Portugal; Department of Pulmonary Disease and Tuberculosis, University Medical Centre Groningen, University of Groningen, Groningen, Netherlands; Tuberculosis Centre Beatrixoord, University Medical Centre Groningen, University of Groningen, Groningen, Netherlands; Mayo Clinic, Pulmonary Disease and Critical Care Medicine, Rochester, MN, USA; Department of Biomedical Sciences, Humanitas University, Via Rita Levi Montalcini 4, 20072, Pieve Emanuele, Milan, Italy, IRCCS Humanitas Research Hospital, Respiratory Unit, Via Manzoni 56, 20089, Rozzano, Milan, Italy; Department of Respiratory Diseases, Amiens-Picardie University Hospital, Amiens, France UR 4294 AGIR, Picardie Jules Verne University, Amiens, France; Division of Mycobacterial and Respiratory Infections, Department of Medicine, National Jewish Health, Denver, Colorado, USA., Division of Infectious Diseases, Department of Medicine, University of Colorado, Aurora, Colorado, USA., Division of Pulmonary Sciences and Critical Care Medicine, Department of Medicine, University of Colorado, Aurora, Colorado, USA; Radboudumc Community for Infectious Diseases, Department of Medical Microbiology, Radboud University Medical Centre, Nijmegen, The Netherlands; Division of Clinical Infectious Diseases, Research Centre Borstel, Borstel, Germany., German Centre for Infection Research (DZIF), Partner Site Borstel-Hamburg-Lübeck-Riems, Borstel, Germany., Respiratory Medicine and International Health, University of Lübeck, Lübeck, Germany. Department of Pediatrics, Global and Immigrant Health, Global Tuberculosis Program, Baylor College of Medicine and Texas Children’s Hospital, Houston, TX, USA; Respiratory Medicine, Royal Free Hospital NHS Foundation Trust, London, & University College London, London UK; Royal Brompton Hospital and NHLI, Imperial College London, London, UK; Clinic for Respiratory Diseases, University Hospital Centre Zagreb, Zagreb, Croatia. University of Zagreb, School of Medicine, Zagreb, Croatia; Respiratory Disease Centre, Fukujuji Hospital, Japan Anti-Tuberculosis Association, Japan. Department of Clinical Mycobacteriosis, Nagasaki University Graduate School of Biomedical Sciences, Japan., Division of Clinical Research, Fukujuji Hospital, Japan Anti-Tuberculosis Association, Japan; Gallipoli Medical Research and Greenslopes Clinical Unit, The University of Queensland, Brisbane Australia; Division of Infectious Diseases, Department of Internal Medicine II, Freiburg University Medical Centre, Freiburg, Germany., Department of Epidemiology, Helmholtz Centre for Infection Research Braunschweig, Germany; Division of Infectious Diseases, Department of Medicine, Oregon Health & Science University, Portland, Oregon, USA., Centre for Infectious Disease Studies, Oregon Health & Science University-Portland State University School of Public Health, Portland, Oregon, USA; Division of Pulmonary and Critical Care Medicine, Department of Internal Medicine, Seoul National University Hospital, Seoul National University College of Medicine, Seoul, Republic of Korea; EPIUnit ITR, Instituto de Saúde Pública da Universidade do Porto, Universidade do Porto, Rua das Taipas, n° 135, 4050-600 Porto, Portugal, Estudo das Populações - Instituto de Ciências Biomédicas Abel Salazar, Universidade do Porto, Porto, Portugal, Centro de Saúde Pública Doutor Gonçalves Ferreira – Instituto de Saúde Pública Doutor Ricardo Jorge, INSA Porto, Porto, Portugal

**Keywords:** Nontuberculous mycobacteria, Treatment resistance, Supportive Care, Antibiotic treatment, Systematic review

## Abstract

**Introduction:** Managing patients with nontuberculous mycobacterial pulmonary disease (NTM-PD) unresponsive to guideline-based therapy presents a significant clinical challenge. Limited treatment options and the complex nature of disease progression make therapeutic decision-making difficult. This study systematically reviewed the existing literature on management strategies and emerging therapeutic approaches for refractory NTM-PD.

**Methods:** We systematically reviewed studies on NTM-PD treatment failure published from inception until 31 January 2024, examining associated factors, intensification strategies, and supportive measures. Twenty-five studies met the inclusion criteria, predominantly exhibiting retrospective observational methodology and focusing on pulmonary disease by *Mycobacterium avium* complex and *Mycobacterium abscessus* species. Findings were synthesised qualitatively due to the considerable heterogeneity in study design and outcomes.

**Results:** Before treatment intensification or de-escalation, the impact of antibiotic treatment on health-related quality of life and microbiological factors, such as acquired resistance and changes in the pathogenic NTM, should be assessed. The presence of severe radiological findings at the beginning of treatment and a history of multiple antibiotic regimen modifications were associated with a lower chance of treatment success. Evidence regarding supportive interventions for treatment-refractory NTM-PD, such as nutritional counselling, respiratory rehabilitation, and psychosocial support, remains limited. Intermittent administration of intravenous antibiotics may be an additional strategy for symptom management. Depending on the causative NTM species, some antibiotic options have been explored for treatment intensification, although most of these are based on observational studies. Surgical resection remains an option for localised disease, although the risk of recurrence remains substantial in the case of preoperative positive culture. Novel therapeutic approaches remain under investigation as potential alternatives.

**Discussion:** This systematic review underscores the complexity of managing refractory NTM-PD, highlighting the role of treatment intensification, symptom control, supportive measures and knowledge gaps. While no standardised approach exists, individualised strategies incorporating clinical, microbiological, and radiological factors remain essential for optimising patient outcomes.

## INTRODUCTION

Nontuberculous mycobacterial pulmonary disease (NTM-PD) is a chronic infection with heterogeneous clinical and microbiological manifestations, associated with significant morbidity and mortality [1–4]. The rising incidence of NTM-PD is attributed to increased life expectancy, a higher prevalence of chronic diseases, widespread immunosuppressive therapy use, and growing awareness of these microorganisms [5–8].

As NTM-PD is diagnosed more frequently, the need for comprehensive and standardised approaches has become evident. Current guidelines were designed to provide therapeutic recommendations for NTM-PD cases based on evidence-based medicine [9, 10]. As such, they cannot offer a structured approach for managing challenges such as treatment-related adverse events, drug intolerance, pathogen re-exposure, symptom management difficulties, and, in particular, treatment failure [9, 10]. Treatment outcomes remain suboptimal, largely due to the complexity of the disease, patients’ frailty, the need for prolonged multidrug antibiotic regimens and ineffective therapies, drug toxicities with poor tolerance and antimicrobial resistance [11, 12]. Despite the established association between NTM-PD and underlying risk factors, particularly bronchiectasis, current therapy recommendations do not include host-directed therapies [9, 10, 13]. Consequently, clinicians often rely on personal expertise and limited available evidence to guide treatment decisions.

This study systematically reviews the available evidence on therapeutic options for patients with NTM-PD refractory to guideline-based therapy (GBT). Our goal was to evaluate alternative approaches and identify gaps in the literature, to further support the development of a structured framework to develop an expert consensus on the management of NTM-PD presenting treatment failure.

## MATERIALS AND METHODS

### Protocol and registration

We performed a systematic review in accordance with the Preferred Reporting Items for Systematic Reviews and Meta-Analyses (PRISMA) guidelines and registered it in the PROSPERO database (PROSPERO 2024. CRD42024491009) [14, 15].

### Search strategy

A comprehensive systematic literature search was performed using PubMed/Medline for studies, concerning human subjects, regarding NTM patients with treatment failure, from inception to 31 January 2024. The search expression was *“nontuberculous mycobacteria AND treatment AND antibiotic*” (Supplementary Material 1 for full query).

### Eligibility and quality assessment

We included case series, cross-sectional, case-control, cohort studies, and clinical trials reporting the management of NTM patients undergoing treatment failure, irrespective of the species. Case reports, conference abstracts, editorials and reviews, and animal or in-vitro studies were excluded. Works in languages other than English were excluded. Treatment failure was defined as persistent culture positivity after six months of GBT, incorporating definitions from Griffith et al. (2018) and the NTM-NET consensus statement (which considered culture positivity after ≥12 months of antimycobacterial therapy) [16, 17].

To address the study’s objectives, we formulated the following research questions:

1. What factors support treatment intensification with curative intent or de-escalation for symptom control in patients with NTM-PD treatment failure?
2. What supportive measures, including non-pharmacological therapies, should be offered?
3. What treatment strategies with curative intent are currently available for treatment intensification?

“Treatment intensification” refers to the modification of the therapeutic regimen to achieve sustained culture conversion and, ultimately, cure, whereas “de-escalation” encompasses treatment halted, as defined by NTM-NET, and the reformulation of the therapeutic plan aiming for symptomatic control, without curative intent [17].

### Data extraction and qualitative assessment

The extracted library was reviewed using EndNote^TM^ 20.2.1 tool (Clarivate, Berkeley, California, United States), by D.MS. and B.M., that were responsible for screening the studies by title and abstract (phase 1) with a third reviewer (A.A.) resolving any eligibility uncertainties in Phase 2 (full-text review). All studies that included data about treatment failure of pulmonary NTM in adult patients were included. Following this, a backward and forward citation assessment was performed.

From each study, the following data was extracted: title, first author; year of publication, the country where the study was conducted; study design, population, and aim; sample size (number of patients included); information on NTM species investigated; treatment outcomes regarding culture sustained conversion rate and treatment period, rate of treatment failure (persistent positive culture), data on clinical improvement, rate of re-infection by other NTM pathogens, and presence of antibiotic resistance.

### Descriptive and qualitative analysis

The authors systematically structured the extracted data from the selected studies to answer the research questions. Given the limited evidence on treatment failure, conducting a meta-analysis was not feasible. Instead, the data were summarised descriptively in tabular format outlining the evaluated interventions and their treatment outcomes. The information on the tables were supplemented with a narrative synthesis of the results and an in-depth discussion, organised by topic, to provide a detailed analysis of the findings and highlight gaps in the literature.

## RESULTS AND DISCUSSION

A total of 5707 studies were retrieved, 135 full-text manuscripts were examined, and 21 studies were included. Additionally, 4 studies were retrieved from citation tracking, making a total of 25 studies included in the review. The flowchart in **Figure 1** synthesises the study selection process.

**Figure 1.**
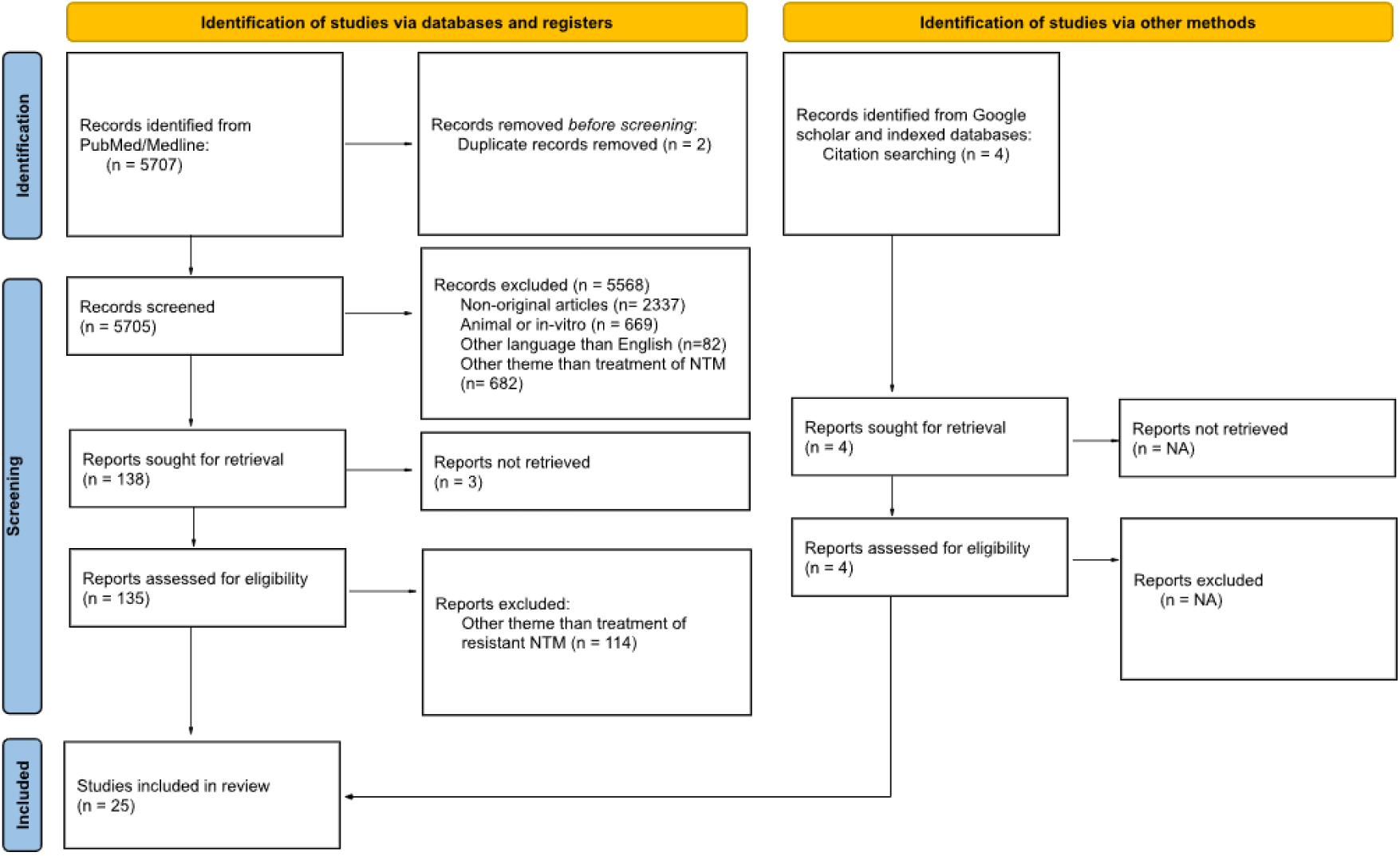
PRISMA flow-chart diagram of included studies.

To assess the quality of the included articles, encompassing 24 non-randomized studies, the authors utilised the validated Mixed Methods Appraisal Tool (MMAT; Version 2018) (Supplementary Material 2) [18]. The majority of the studies did not meet the criteria for accounting for confounders in the analysis.

With the available evidence, seven major topics were identified: Quality of Life and Symptom Management, Radiological Severity and Treatment Failure, Microbiological Factors and Resistance Patterns, Impact of Prolonged Antibiotic Regimens, Supportive Measures and Non-Pharmacological Approaches, Intermittent Antibiotic Therapy for Symptom Control and, Alternative Treatment Strategies After Guideline-Based Treatment Failure.

From the 25 studies included (Table 1), most were retrospective cohort studies (n=17). The research focused on *Mycobacterium avium* complex (MAC) (n=16) and/or *Mycobacterium abscessus* (MABS) pulmonary disease (n=17) [16, 19–42]. The studies varied in design, patient populations, microbiological endpoint definitions, and treatment regimens, limiting direct comparability. While none specifically examined clinical decision-making following treatment failure, some identified factors linked to treatment response, which could have implications for patient management (Table 1).

**Table 1.**
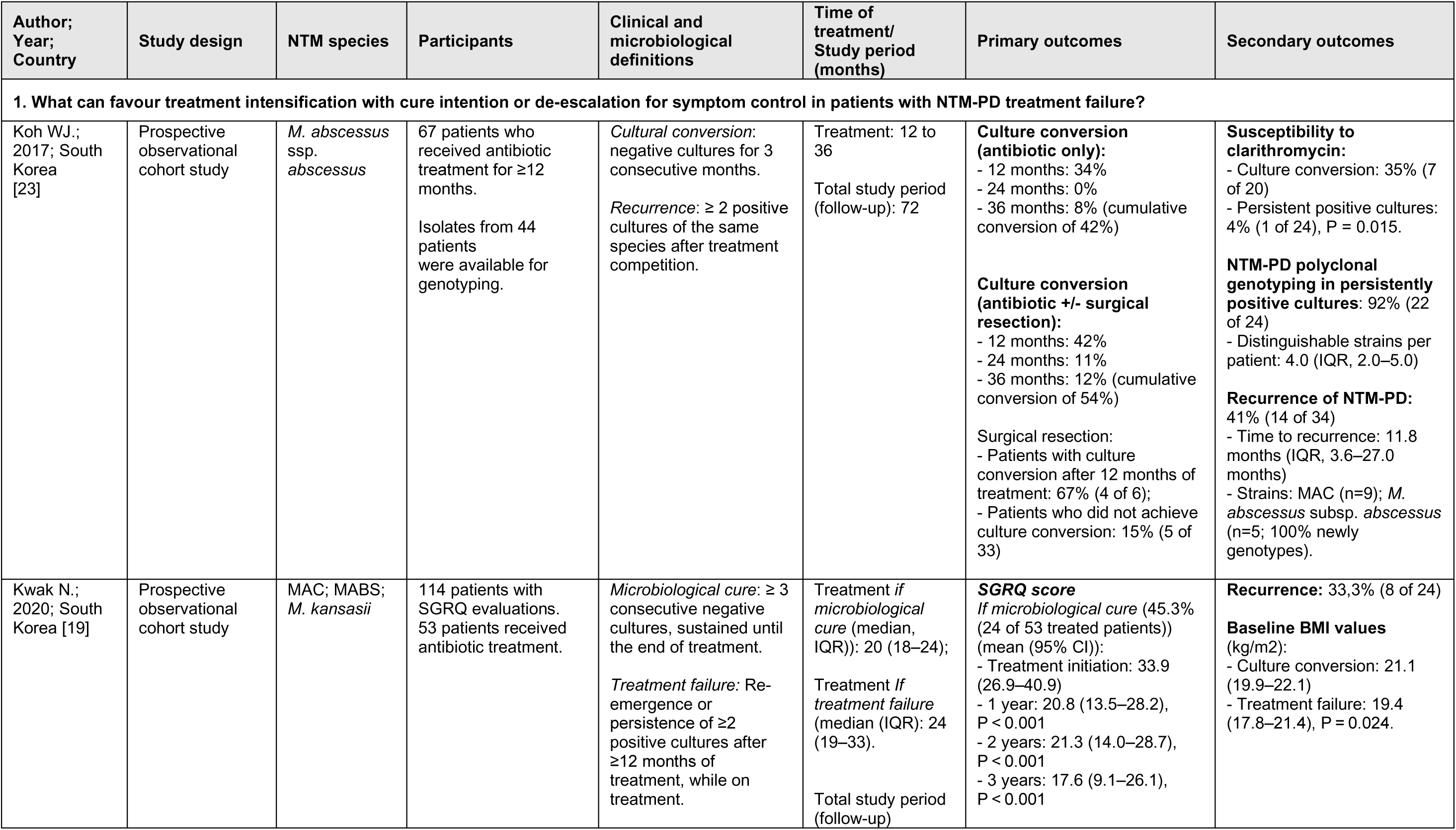
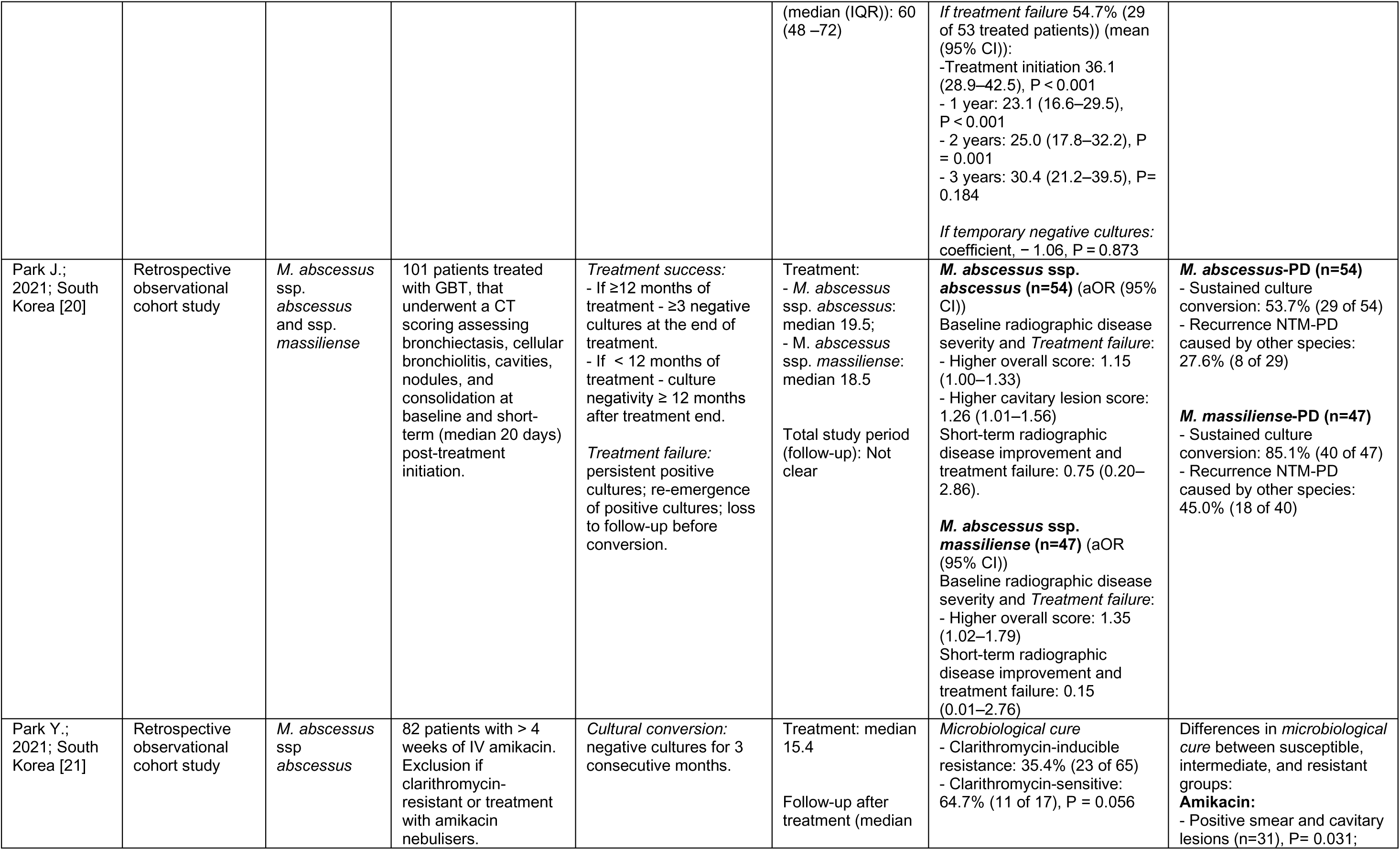
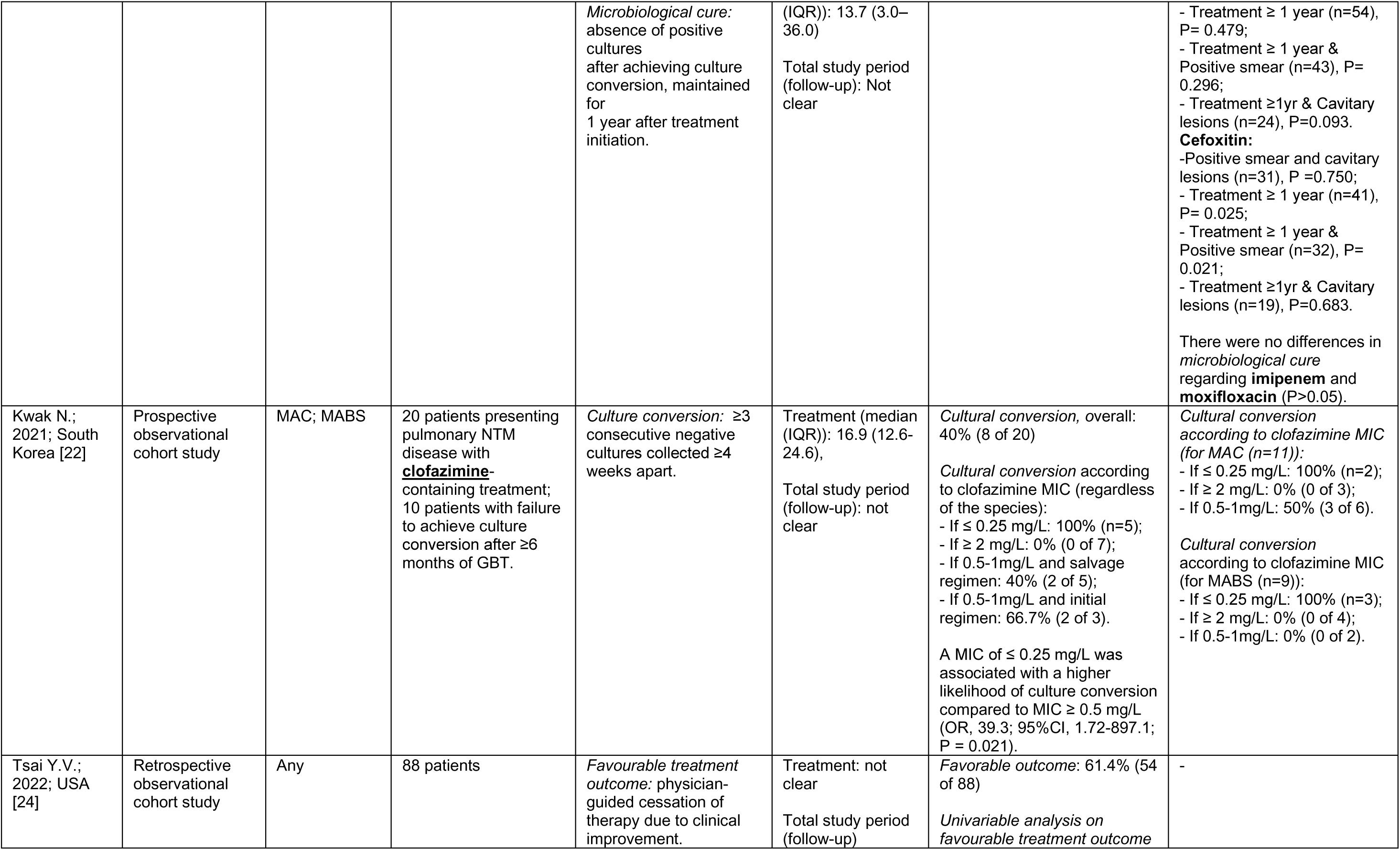
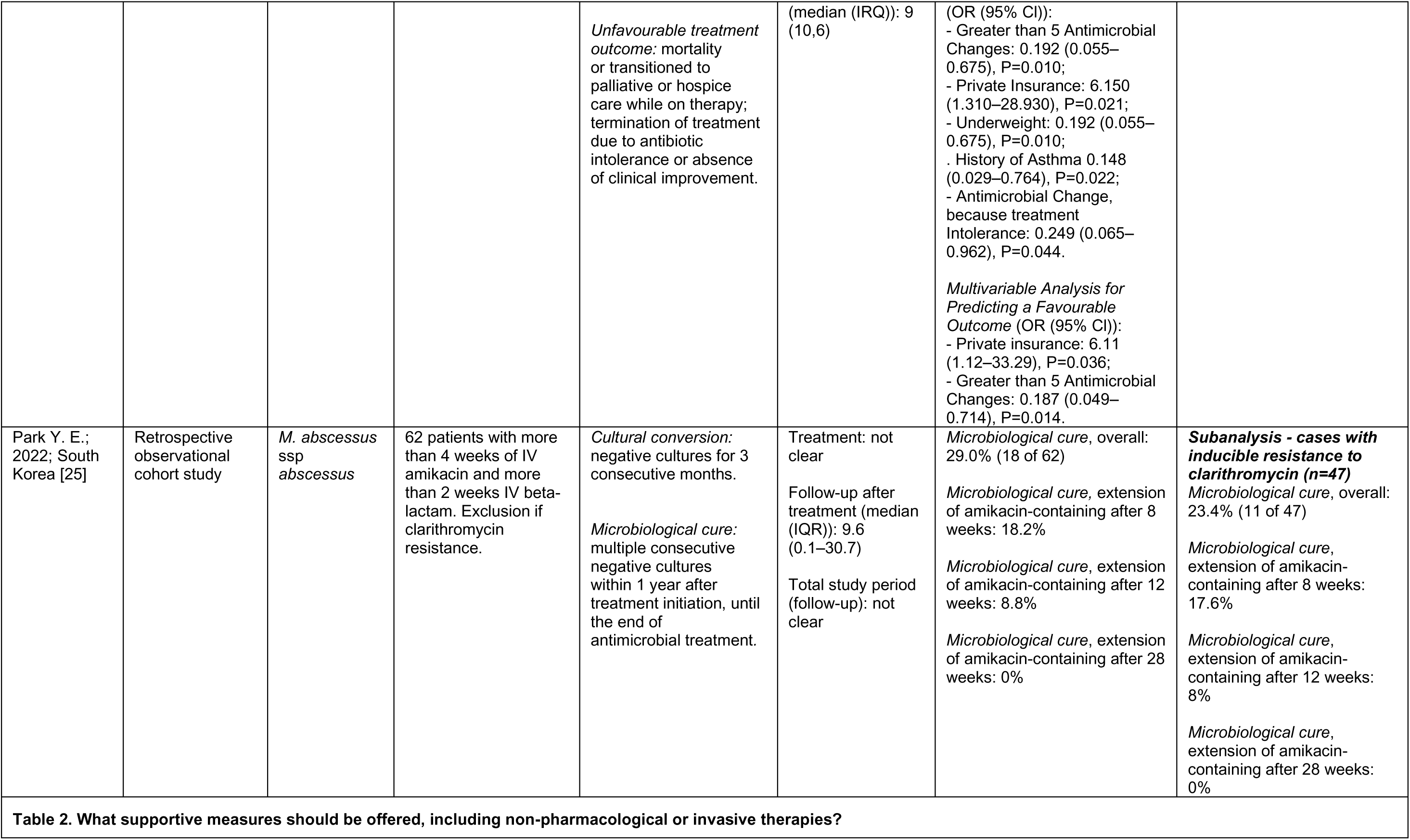
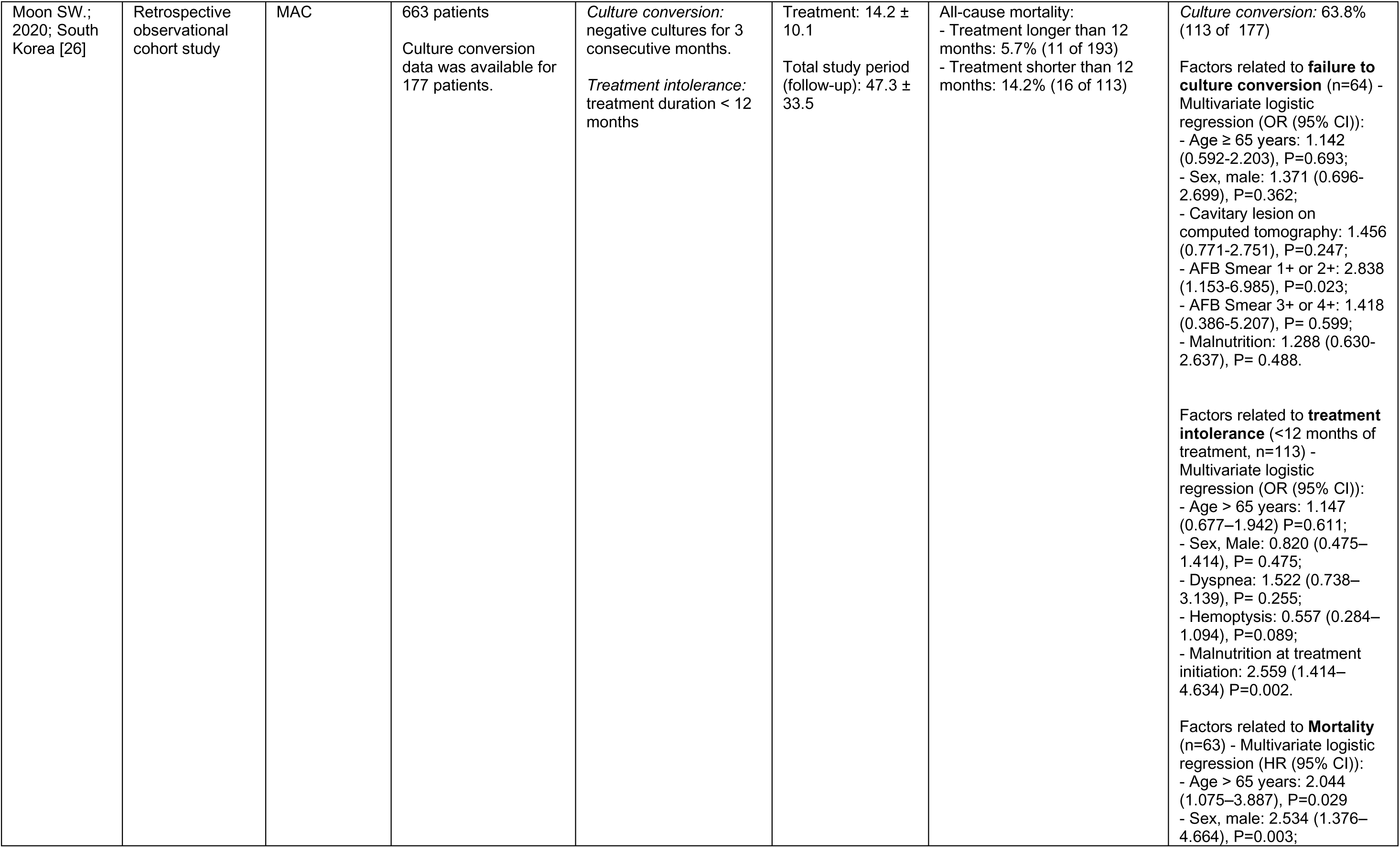
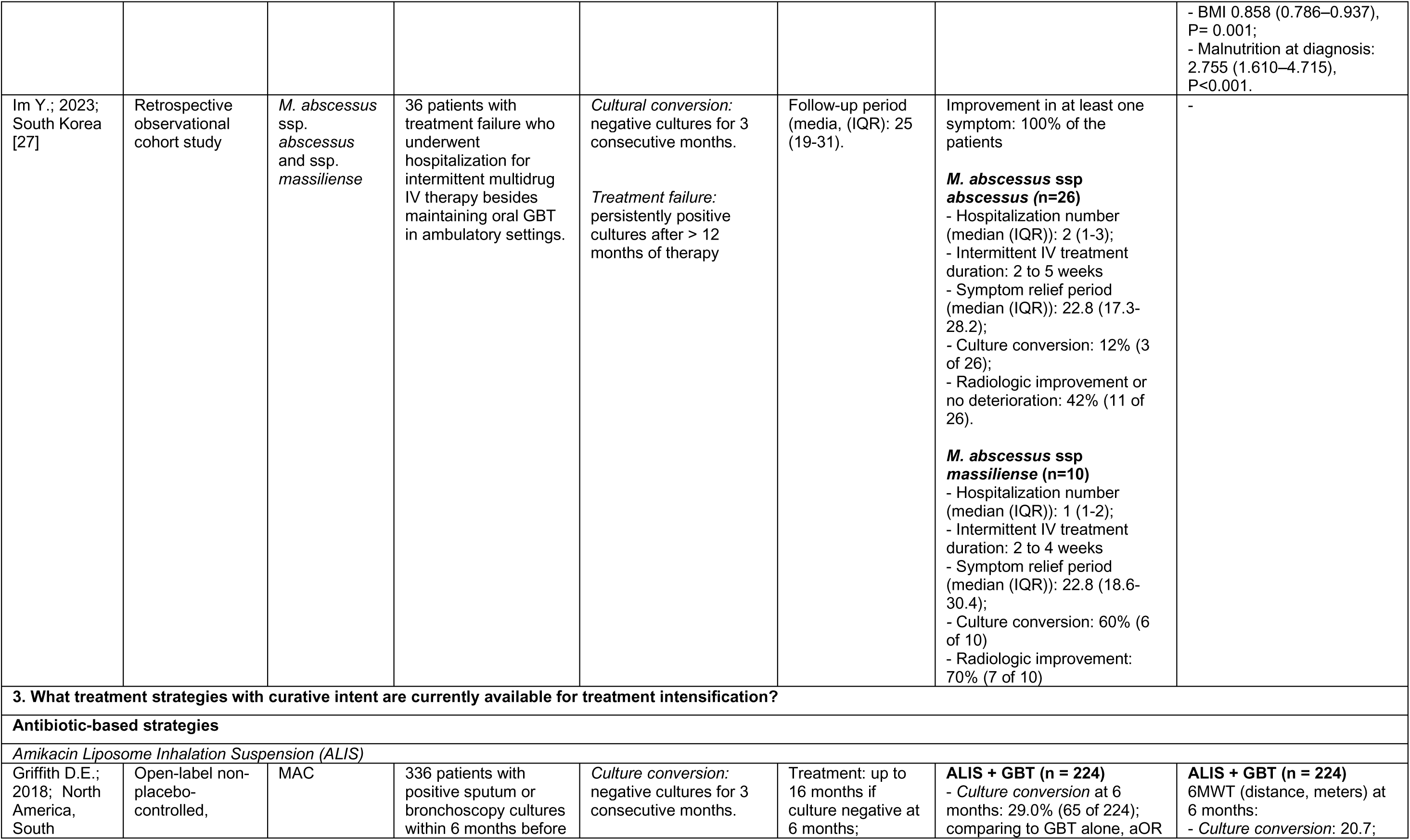
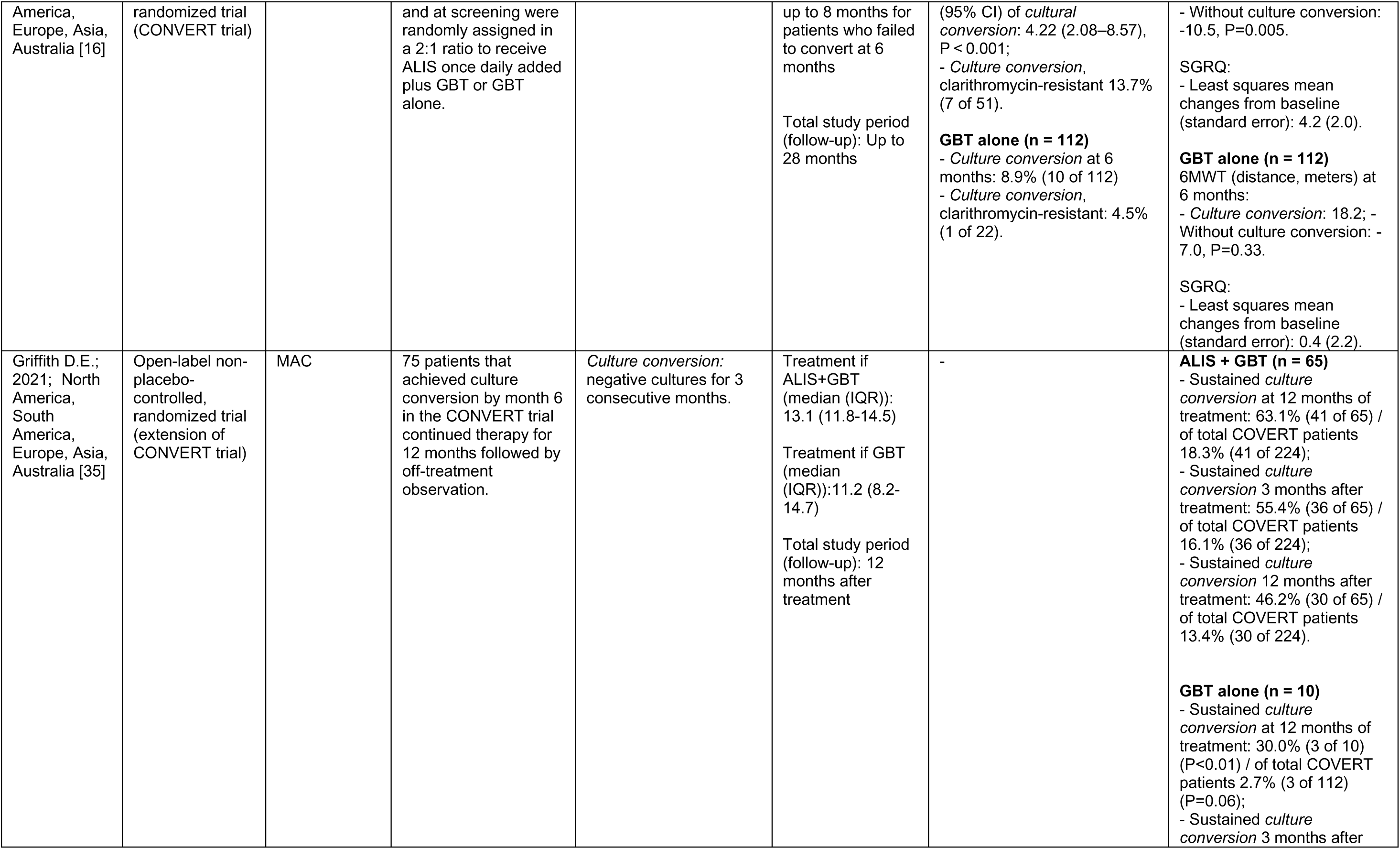
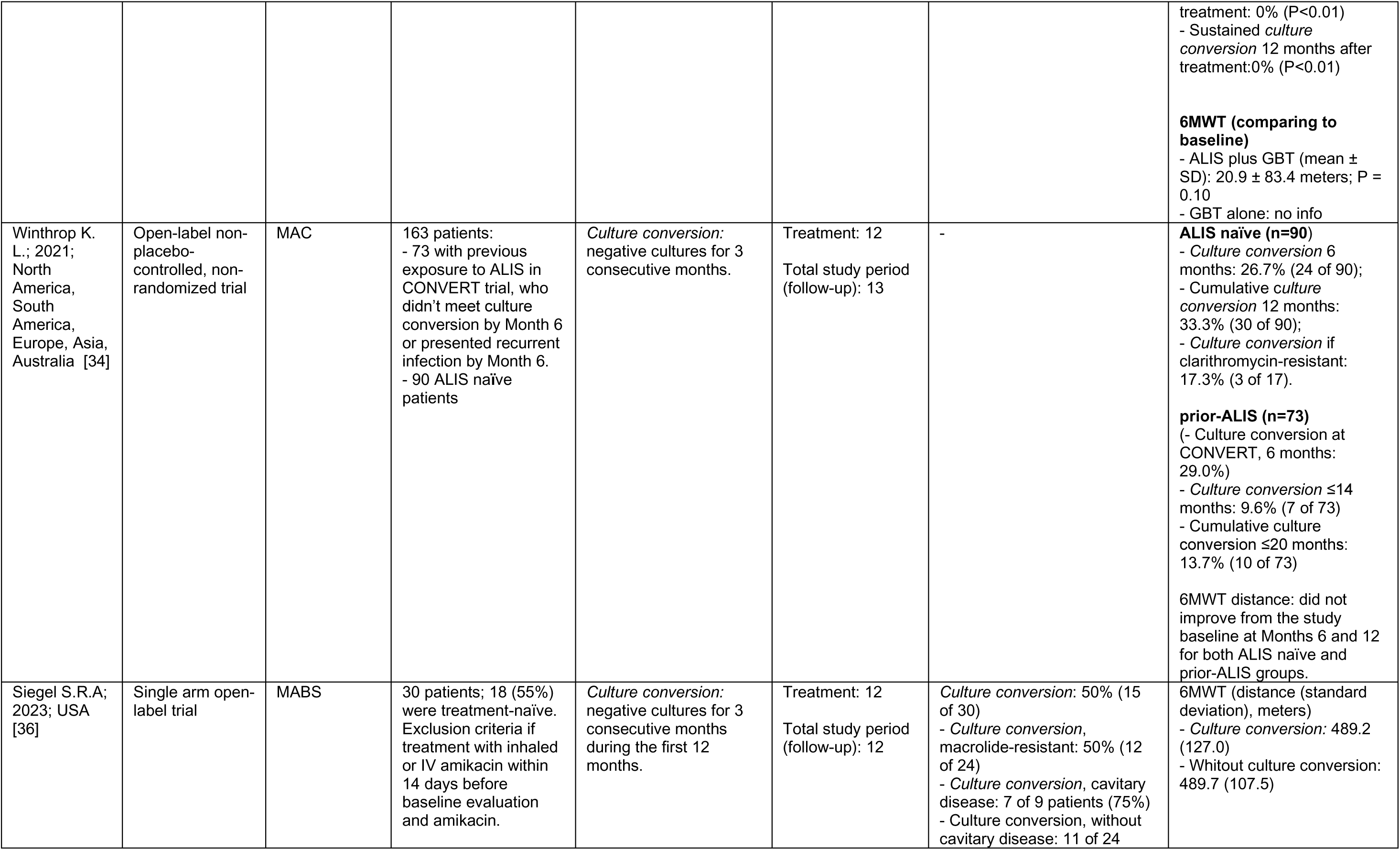
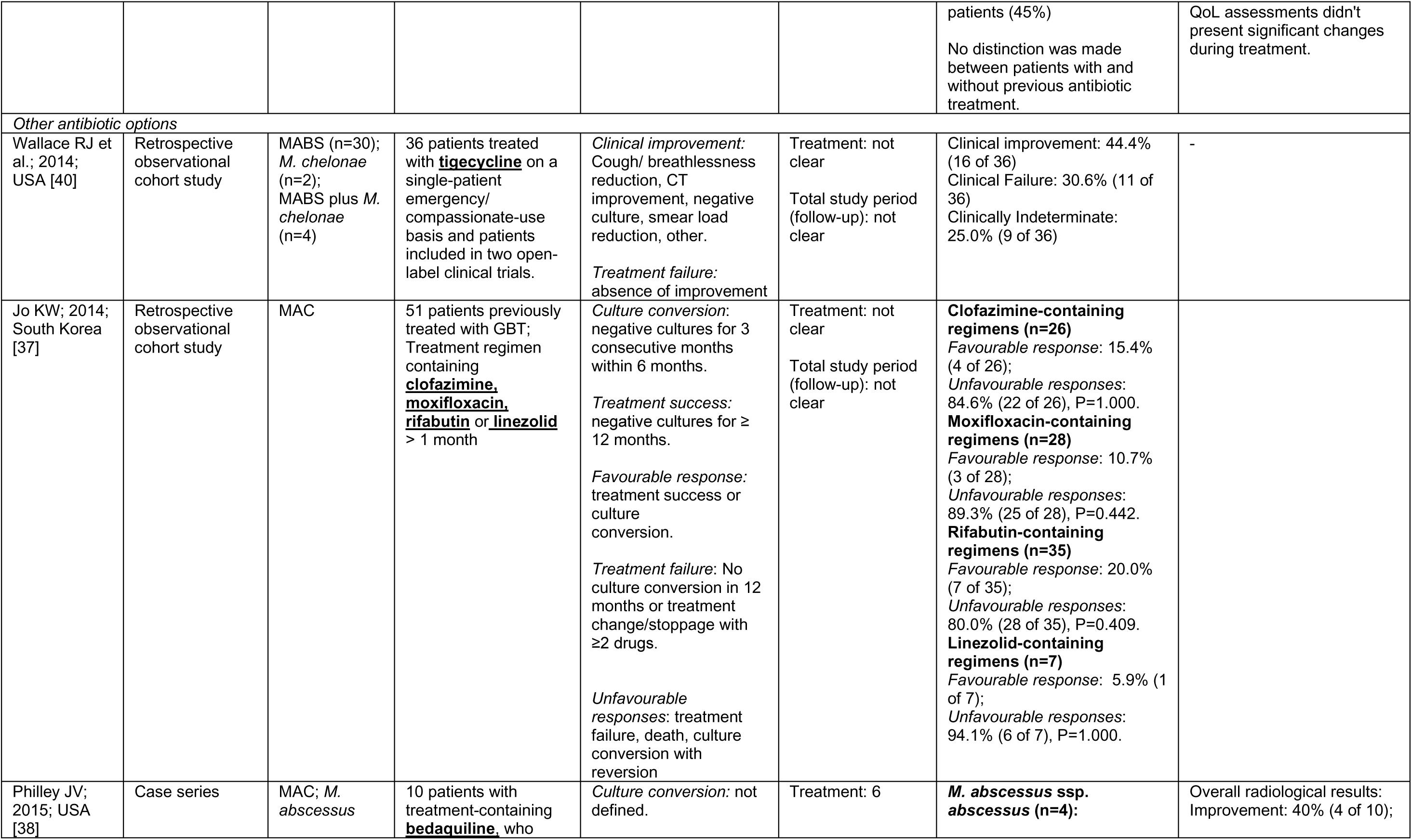
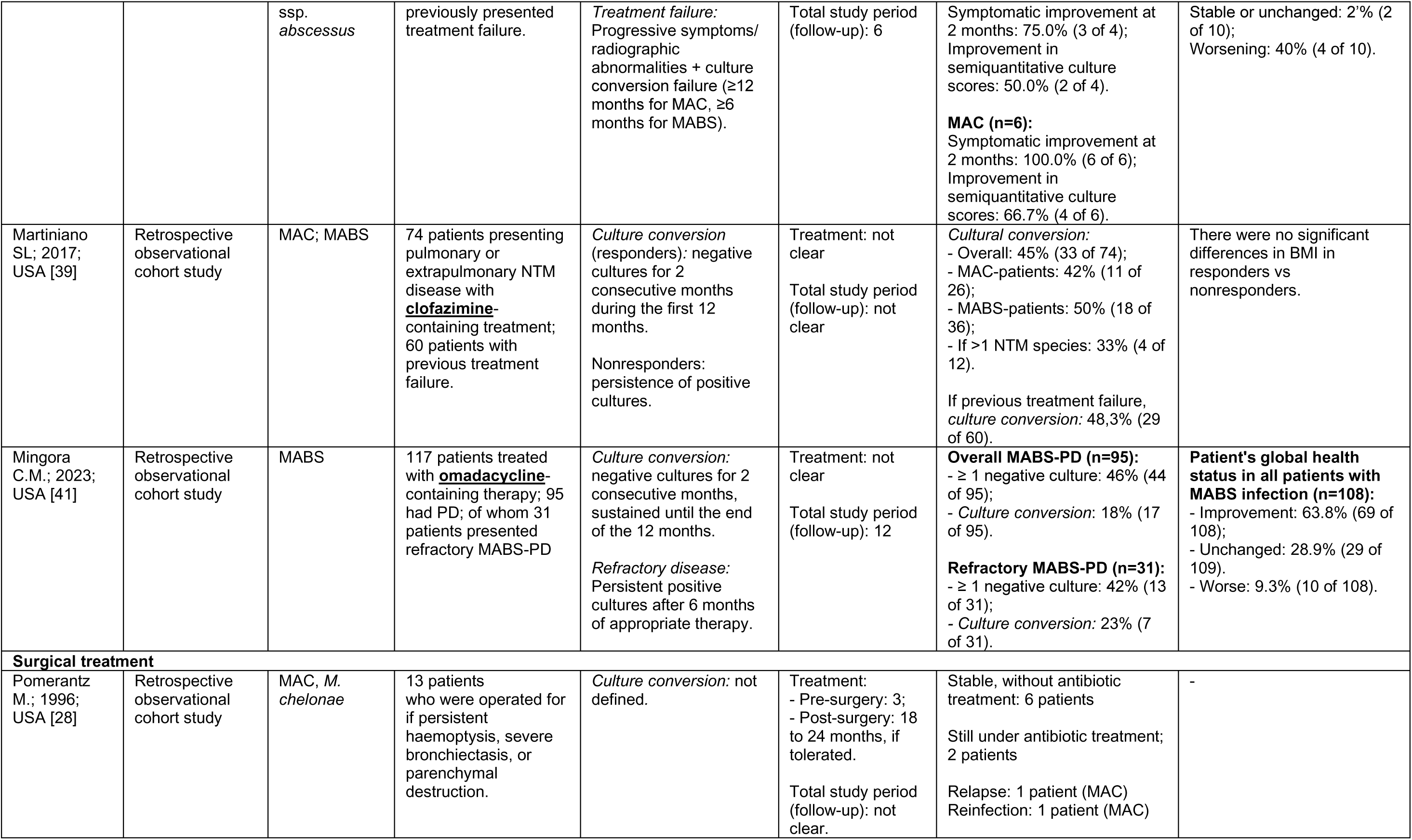
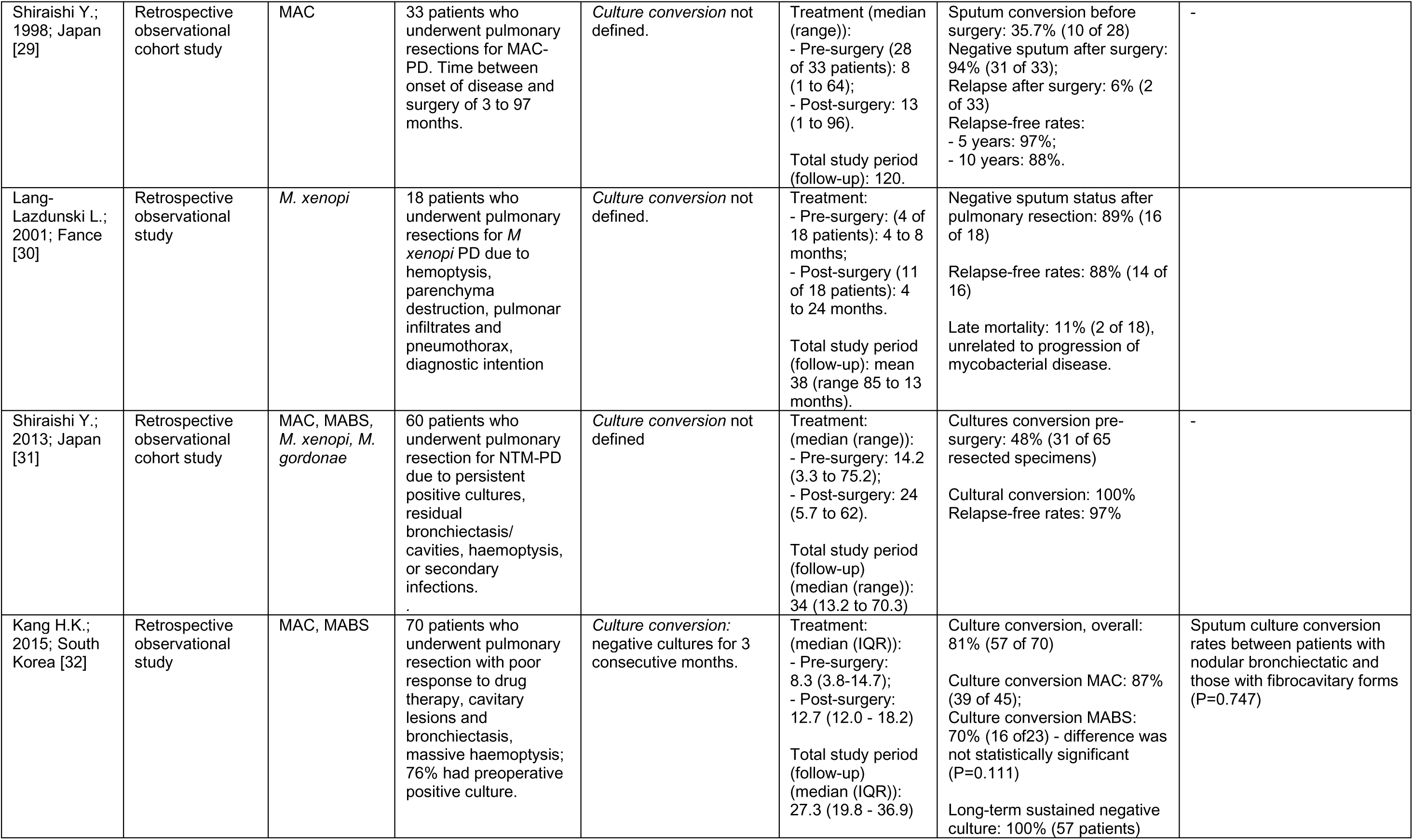
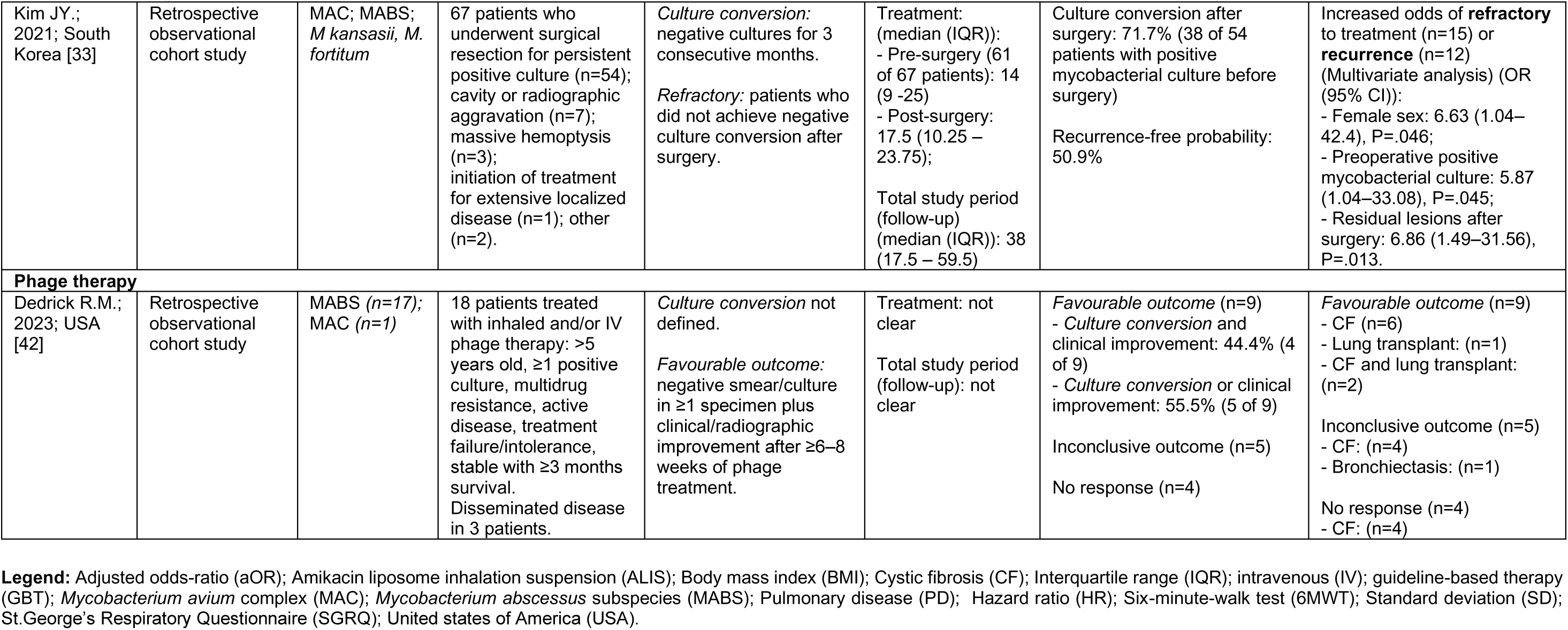
Characteristics of the selected articles.

### Quality of Life (QoL) and Symptom Management

Kwak et al. (2020) assessed health-related QoL changes by applying the St. George’s Respiratory Questionnaire (SGRQ) in 114 NTM-PD patients, of whom 53 received antibiotic treatment and 24 achieved microbiological cure [19]. The study revealed that QoL improved within the first two years of treatment, even among those classified as treatment failures (n=29), with sustained improvements observed at three years. Whether these changes in QoL correlate with bacterial load or other disease parameters remains uncertain. However, these findings indicate that antibiotic treatment may be associated with symptomatic improvement, despite persistent positive cultures.

### Radiological Severity and Treatment Failure

Radiological severity has been proposed as a potential predictor of treatment failure. Park et al. (2021) observed that a higher radiological disease burden at the initiation of antibiotic treatment was associated with an increased risk of treatment failure in MABS-PD [20]. However, short-term radiographic changes (after a median of 20 days) showed no association with treatment outcomes. Kim et al. (2024) recently reported findings from a cohort of MAC-PD patients, showing that post-treatment radiographic severity and its improvement from baseline after 12 months of treatment were associated to a reduction in all-cause mortality [43]. This association was observed regardless of culture conversion status, as the study found no significant correlation between achieving culture conversion and severity of radiographic findings. The utility of radiographic severity scores in guiding treatment adjustments after initial treatment failure remains uncertain. Additionally, radiological evaluation is challenging due to underlying lung structural disease and immunosuppression, where deterioration may be linked to co-infections or other pathogens [44–46].

### Microbiological Factors and Resistance Patterns

Macrolide and amikacin resistance in MAC and MABS-PD is associated with an increased likelihood of treatment failure, aligning with the current understanding of NTM-PD treatment challenges [9, 10, 16, 34, 47]. For antimicrobials other than macrolides and amikacin, the absence of clinically validated breakpoints complicates associating *in vitro* susceptibility and clinical response [48]. Park et al. (2021) found significant differences in microbiological outcomes only in subgroups of patients with cefoxitin and amikacin susceptibility in a cohort of*. M. abscessus* ssp. *abscessus* PD patients [21]. Similarly, Kwak et al. (2021) identified an association between negative sputum culture conversion and lower minimum inhibitory concentrations (MICs) for clofazimine in patients with MAC and MABS-PD, although standardised cutoff values have yet to be established [22].

Additionally, genotypic variation during treatment and at relapse has been previously described in works with MAC-PD cohorts [49, 50]. A study with a cohort of patients *M. abscessus* ssp. *abscessus* PD under treatment for ≥ 12 months identified genotypic variation in 92% of the cases with persistently positive cultures during antibiotic treatment and in all microbiologic recurrences post-treatment [23]. Additionally, an in-vitro and in-animal recent study demonstrated that M. avium presents intrinsic heteroresistance to clarithromycin in vivo, which may justify inadequate treatment response in cases with susceptible antibiotic susceptibility test (AST) results [51]. The clinical implications of both genotyping variation and NTM strains’ intrinsic heteroresistance require further investigation.

### Impact of Prolonged Antibiotic Regimens

One of the challenges in managing NTM infections is the prolonged course of antibiotics, which is often associated with adverse effects and difficulties in treatment adherence [52]. Tsai et al. (2022) reported an association of a threshold of more than five changes to antimicrobial regimens with an 81.3% increased risk of poor outcomes, including mortality, treatment discontinuation, or intolerance [24]. These findings suggest that frequent modifications to regimens, often justified by intolerance or failure to respond, are linked to worse clinical outcomes, although the causality of this relationship was only established in this study. Conversely, private insurance was associated with more favourable clinical outcomes, which was attributed by the authors to enhanced access to more treatment options, suggesting that the healthcare system may also have an impact on the outcome [24].

Another study of MABS-PD patients revealed that the administration of intravenous amikacin beyond 28 weeks did not result in the achievement of additional sustained negative cultures, thus demonstrating poor outcomes for patients receiving regimens with extended amikacin periods beyond the guideline 16-week recommendation [25]. These results underscore the need for careful treatment planning, balancing regimen modifications with patient tolerance, and the likelihood of successful outcomes. Further research is required to refine treatment strategies, optimise long-term management and define the optimal treatment duration for these patients.

Given the challenges associated with prolonged antibiotic regimens, exploring supportive measures and non-pharmacological approaches becomes essential in improving patient outcomes and treatment tolerance.

### Supportive Measures and Non-Pharmacological Approaches

The role of non-pharmacological interventions in NTM-PD treatment failure remains inadequately defined. Some studies have examined nutritional status, rehabilitation programmes, and the psychological burden of the disease, but the evidence on effective interventions continues to be limited [26, 53–56]. Moon et al. (2020) found that malnutrition was not associated with culture conversion failure but was linked to higher mortality and increased treatment intolerance [26]. However, in this study, the influence of nutritional advice on clinical or microbiological outcomes was not assessed. The only cohort study on respiratory rehabilitation in NTM-PD focused on untreated patients and reported improvements in cough, sputum production, functional status, and radiological findings in some cases [55]. Chest physiotherapy combined with hypertonic saline inhalation has been investigated as a supportive measure, though the available data remains limited to one study [57]. Currently, non-pharmacological and symptomatic interventions are guided by expert opinion and extrapolation from other respiratory diseases, such as bronchiectasis [13, 58].

### Intermittent Antibiotic Therapy for Symptom Control

Intermittent antibiotic regimens have been explored to reduce treatment burden in patients experiencing intolerance to long-term therapy. Im et al. (2023) reported that intermittent IV antibiotic courses provided symptom relief for a median duration of 22.8 months in MABS-PD patients [27]. Although it is important to highlight that there are no data on patient outcomes following the complete discontinuation of antibiotics, making it unclear which patients may benefit from this approach, these findings suggest that anticipated planning of antibiotic regimens that allow symptom-suppressive intermittent phases may be advantageous in cases of treatment-resistant microbiological conditions.

### Alternative Treatment Strategies After Guideline-Based Treatment Failure

Alternative strategies for NTM-PD treatment failure include surgical interventions, modifications in antibiotic regimens, and experimental therapies [16, 28–42].

Surgery has been evaluated in localised NTM-PD cases, particularly those with structural lung disease or haemoptysis. The frequency of reported culture conversion ranges between 71.7% and 94.0% [28–33]. However, the presence of residual bronchiectasis, contralateral nodules, and, most importantly, preoperative positive cultures is associated with higher postoperative recurrence rates, making surgery a less reliable option [32].

Studies on antibiotic-based treatment intensification have primarily focused on Amikacin Liposome Inhalation Suspension (ALIS), which has been shown to improve culture conversion rates in MAC-PD and was therefore incorporated into the GBT [9]. In fact, ALIS has been associated with a 29.0% culture conversion rate at 6 months in treatment-refractory MAC-PD, with a cumulative conversion of up to 33.3% after 12 months, and sustained culture conversion in 46.2% of the converter patients 12 months after treatment [16, 34, 35]. The effectiveness of ALIS in MABS-PD remains less well-established, though preliminary data suggest a potential microbiological benefit [36].

Additionally, limited data are available on the use of other agents, including clofazimine, bedaquiline, tigecycline, and omadacycline, highlighting the need for further research on their role in treatment optimisation.

Regarding MAC-PD, clofazimine and bedaquiline lack robust evidence to incorporate treatment schemes in cases of treatment failure [37–39]. However, clofazimine has been frequently studied in treatment-naïve cohorts or as a component of alternative regimens, showing more promising results [59–63], and inclusion of bedaquiline in the treatment scheme for treatment-refractory MAC-PD is the subject of an ongoing clinical trial (ClinicalTrials.gov identifier NCT04630145 [64]). No substantial evidence supports other antibiotic options’ efficacy, including moxifloxacin, rifabutin, or linezolid [37].

For MABS-PD, antibiotic approaches studied for treatment failure are already included as first-line options in NTM guidelines, including clofazimine and 3rd-generation tetracyclines, as omadacycline and tigecycline [9, 10]. Observational studies have demonstrated the potential efficacy of these antibiotics when utilised as additional components of treatment regimens or as alternative pharmacological options [38–41, 65, 66]. The current evidence does not allow for a definitive conclusion regarding which approach must be considered, as decisions were tailored case-by-case. Furthermore, ongoing investigations on novel antibiotics for refractory MABS-PD, including Delpazolid, may expand therapeutic options (ClinicalTrials.gov identifier NCT06004037 [67]).

Phage therapy has been explored as a potential treatment option for refractory MABS-PD, with a preliminary case series suggesting microbiological and clinical improvements in some patients [42]. However, the long-term impact and optimal patient selection criteria remain unclear, and further results of clinical trials are required (ClinicalTrials.gov identifier NCT06262282 [68]). Additional studies have explored the potential role of host-directed therapies in modifying the immune response and underlying comorbidities, though further supporting data are needed [69, 70].

### Strengths and Limitations

This systematic review possesses several strengths, including adherence to PRISMA guidelines and registration with PROSPERO, which ensures methodological rigour. A comprehensive search strategy minimised selection bias, and the focus on treatment failure in NTM-PD addresses a critical gap in the literature. Unlike previous reviews, this study assesses both treatment intensification and de-escalation, integrating pharmacological and non-pharmacological approaches in patients with refractory NTM-PD. Moreover, the incorporation of symptomatic control and quality of life (QoL) enhancement as treatment objectives provides an opportunity to reflect upon existing therapeutic approaches that focus on microbiological outcomes.

However, this review also has limitations. The heterogeneity of study designs, populations, and treatment regimens precluded a statistical analysis. Most of the studies included were small retrospective cohorts, increasing the risk of bias. The lack of standardised endpoints complicated comparisons, and long-term outcomes remain largely underexplored. Non-pharmacological interventions were underrepresented, which limits insights into supportive care strategies. Potential publication bias may also have led to the overrepresentation of successful interventions.

### Summary of Findings

This systematic review highlights significant gaps in the management of NTM-PD treatment failure, aiming to identify key areas requiring further evidence and to support the development of expert consensus. Several factors, including QoL score changes, radiological severity, resistance patterns, and treatment prolongation, were examined but none offered a clear decision-making framework. The role of non-pharmacological interventions, alternative therapies, and emerging treatments remains poorly defined, with limited evidence on long-term outcomes. Further research is needed to clarify the clinical utility of different approaches, particularly in guiding treatment adjustments after GBT failure. Establishing a standardised definition of treatment success and distinguishing between reinfection and treatment failure through genotypic identification of NTM species could enhance the evaluation of treatment regimens in the future.

## Data Availability

All data produced in the present work are contained in the manuscript

