## Supplementary material S1. for "Management of Nontuberculous Mycobacterial Pulmonary Disease Refractory to Guideline-Based Therapy: A Systematic Review"

**Supplementary material S1.** Full query used for the systematic review

("nontuberculous mycobacteria"[MeSH Terms] OR ("nontuberculous"[All Fields] AND "mycobacteria"[All Fields]) OR "nontuberculous mycobacteria"[All Fields]) AND ("therapeutics"[MeSH Terms] OR "therapeutics"[All Fields] OR "treatments"[All Fields] OR "therapy"[MeSH Subheading] OR "therapy"[All Fields] OR "treatment"[All Fields] OR "treatment s"[All Fields]) AND ("anti bacterial agents"[Pharmacological Action] OR "anti bacterial agents"[MeSH Terms] OR ("anti bacterial"[All Fields] AND "agents"[All Fields]) OR "anti bacterial agents"[All Fields] OR "antibiotic"[All Fields] OR "antibiotics"[All Fields] OR "antibiotic s"[All Fields] OR "antibiotical"[All Fields])
