## Supplementary material S2. for "Management of Nontuberculous Mycobacterial Pulmonary Disease Refractory to Guideline-Based Therapy: A Systematic Review"

**Supplementary material S2.** Quality assessment according to the Mixed Methods Appraisal Tool.

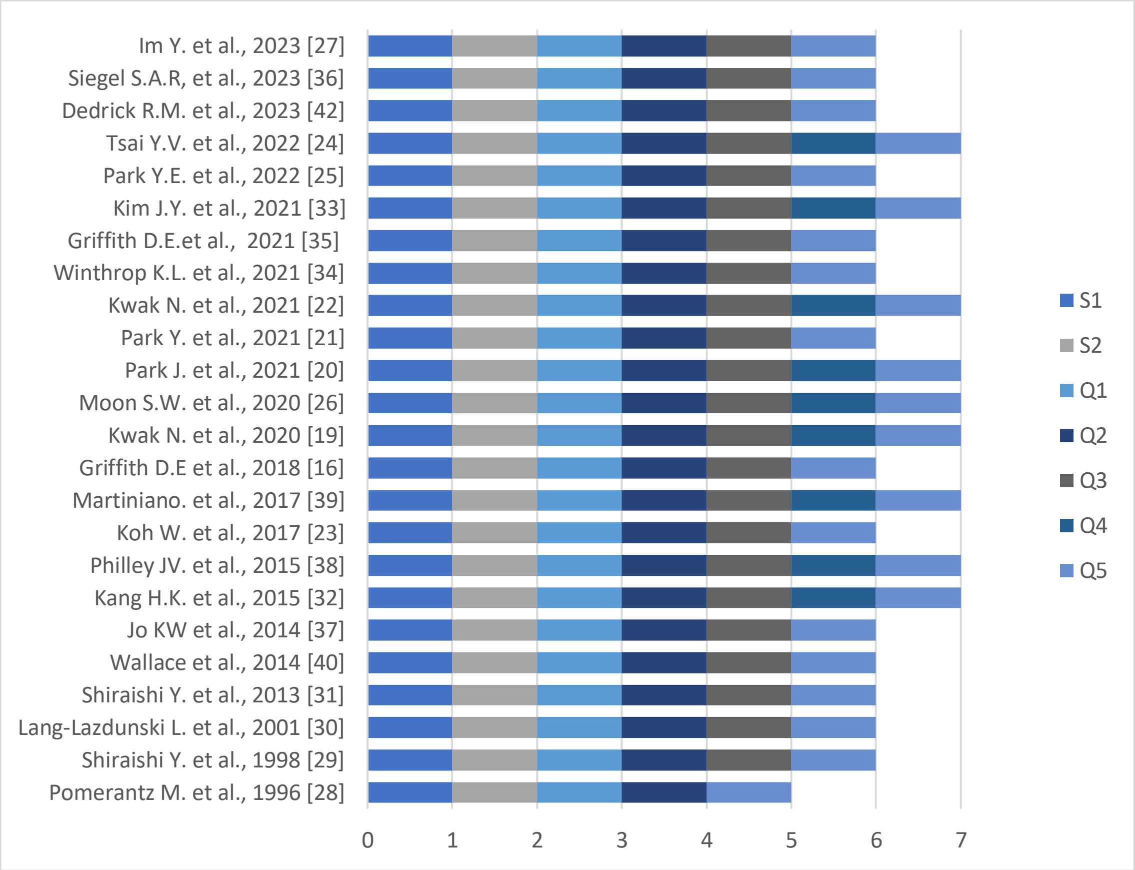


According to the MMAT tool, each included study was classified into an appropriate design category. The two initial screening questions were equal for all the studies (*S1. Are there clear research questions? S2. Do the collected data allow me to address the research questions?).* The qualitative questions vary between different study designs. This systematic review included: **Quantitative randomized controlled trials** (*1. Is randomization appropriately performed? 2. Are the groups comparable at baseline? 3. Are there complete outcome data? 4. Are outcome assessors blinded to the intervention provided? 5. Did the participants adhere to the assigned intervention?);* **Quantitative non-randomized studies** (*1. Are the participants representative of the target population? 2. Are measurements appropriate regarding both the outcome and intervention (or exposure)? 3. Are there complete outcome data? 4. Are the confounders accounted for in the design and analysis? 5. During the study period, is the intervention administered (or exposure occurred) as intended?).* Based on the quality assessment, there were no studies excluded; and **Qualitative studies**: (*1. Is the qualitative approach appropriate to answer the research question? 2. Are the qualitative data collection methods adequate to address the research question? 3. Are the findings adequately derived from the data? 4. Is the interpretation of results sufficiently substantiated by data? 5. Is there coherence between qualitative data sources, collection, analysis and interpretation?).* List of abbreviations: Y (yes), N (no).
